# Chronic Kidney Disease Amplifies Gabapentin-Associated Dementia Risk in Non-Dialysis Patients: Findings Consistent with a Renal Pharmacokinetic Vulnerability

**DOI:** 10.64898/2026.03.15.26348418

**Authors:** James Green, Laura D. Byham-Gray, Joshua Kaplan, Suril Gohel, Branimir Ljubic, Michael Schulewski, Barbara Tafuto

**Affiliations:** Department of Health Informatics, Rutgers School of Health Professions, Newark, New Jersey; Department of Clinical and Preventive Nutrition Sciences, Rutgers School of Health Professions, Newark, New Jersey; Division of Nephrology, Department of Medicine, Rutgers New Jersey Medical School, Newark, New Jersey; New Jersey Alliance for Clinical and Translational Science, Rutgers University, New Brunswick, New Jersey; Office of Advanced Research Computing (OARC), Rutgers University, Piscataway, New Jersey

**Keywords:** chronic kidney disease, gabapentin, pregabalin, dementia, pharmacoepidemiology, drug safety, renal pharmacokinetics

## Abstract

Gabapentinoids are widely prescribed in patients with chronic kidney disease (CKD), yet whether routine renal dose adjustment is sufficient to mitigate cognitive safety risk remains unresolved. Gabapentin and pregabalin differ markedly in pharmacokinetic behavior under impaired clearance: gabapentin half-life extends from 5–7 hours to 52–132 hours in advanced CKD (a 10-to 20-fold increase), versus an approximately 4-fold extension for pregabalin. We examined whether CKD amplifies gabapentin-associated dementia risk relative to pregabalin in a real-world active comparator cohort, with external replication.

Among 33,791 adults aged ≥40 years with hypertension initiating gabapentinoids in the Rutgers Clinical Research Data Warehouse (2018–2024), gabapentin was associated with substantially elevated dementia risk in patients with CKD (hazard ratio [HR] 7.39; 95% CI, 3.43–15.92; *P*<0.001) versus a near-null association in patients without CKD (HR 1.09; 95% CI, 0.89–1.34; *P*=0.41). The elevated risk in CKD persisted within the low-dose stratum (≤300 mg: HR 5.06 in CKD vs. 1.27 in non-CKD), suggesting that dose adjustment alone may not fully offset cognitive safety risk. The signal concentrated in non-dialysis CKD (KDIGO G3b–G4: HR 4.54; 95% CI, 1.62–12.76) and attenuated in CKD stage 5 (HR 1.57; *P*=0.18), a pattern consistent with renal pharmacokinetic exposure as the dominant driver. External replication in the NIH All of Us Research Program (N=47,079) confirmed the gabapentin–pregabalin signal (HR 1.59; 95% CI, 1.35–1.88; *P*<0.001) with a directionally consistent eGFR gradient. FAERS pharmacovigilance analysis showed disproportionately higher renal adverse event reporting for gabapentin versus pregabalin (chronic kidney disease ROR 5.98; renal failure ROR 2.43).

These findings are consistent with the interpretation that CKD may transform routine gabapentin prescribing into a renal pharmacokinetic cognitive safety problem, with risk concentrated in the non-dialysis CKD population and persisting even at low prescribed doses. Standard dose adjustment alone may be insufficient. Renal function stage warrants closer integration into gabapentinoid selection in patients with CKD, with pregabalin a candidate alternative where cardiovascular risk permits.

**Key Points:**

- In an active comparator new-user cohort of 33,791 gabapentinoid initiators, gabapentin was associated with substantially elevated dementia risk in patients with chronic kidney disease (CKD) compared with pregabalin, with the excess risk persisting even within the low-dose stratum (≤300 mg) — a pattern consistent with the interpretation that standard renal dose adjustment alone may not fully offset the cognitive safety risk of gabapentin in CKD.
- The signal concentrated in non-dialysis CKD (KDIGO stages G3b–G4) and attenuated in CKD stage 5, paralleling expected pharmacokinetic exposure: gabapentin half-life extends 10-to 20-fold in advanced CKD versus approximately 4-fold for pregabalin, while dialysis partially restores clearance.
- Convergent evidence from external replication in the NIH All of Us Research Program (N=47,079) and FDA FAERS pharmacovigilance data — including disproportionately higher renal adverse event reporting for gabapentin versus pregabalin (chronic kidney disease reporting odds ratio [ROR] 5.98; renal failure ROR 2.43) — supported the interpretation of a renal pharmacokinetic cognitive safety vulnerability.
- These findings suggest that renal function stage warrants closer integration into gabapentinoid selection in older adults with CKD, with pregabalin a candidate alternative where cardiovascular risk permits.

## 1. Introduction

Gabapentinoids — gabapentin and pregabalin — are among the most commonly prescribed medications in patients with chronic kidney disease (CKD), where they are used for diabetic and uremic peripheral neuropathy, restless legs syndrome, neuropathic pain, and pruritus.^1,2^,^11,12,13^ Both agents are renally eliminated and require dose reduction as renal function declines, but their pharmacokinetic behavior under impaired clearance is markedly different. In advanced CKD, gabapentin half-life extends from 5–7 hours to approximately 52–132 hours — a 10-to 20-fold increase — while pregabalin half-life extends approximately 4-fold.^6^ Gabapentin also exhibits saturable, nonlinear absorption, with bioavailability falling from roughly 60% to 27% as oral dose increases, producing erratic plasma concentrations even at standard doses.^3^ Pregabalin, by contrast, maintains linear pharmacokinetics and >90% bioavailability across the clinical dose range.^4,5^ These differences imply that, in the same CKD patient, gabapentin and pregabalin may produce substantially different central nervous system (CNS) exposure profiles even when both are dose-adjusted to standard guideline recommendations.

Whether this differential pharmacokinetic vulnerability translates into a cognitive safety signal that persists despite renal dose adjustment is not well characterized. Most published pharmacoepidemiologic analyses of gabapentinoids and dementia have used non-user comparators and have not stratified by renal function,^7,8,9^ which can dilute a signal that concentrates in the CKD population. At the same time, gabapentinoids are among the medications most commonly associated with adverse events in dialysis patients.^13^

We therefore examined whether CKD amplifies the dementia risk associated with gabapentin compared with pregabalin in a real-world active comparator new-user cohort, and whether any such signal is reproducible across independent data sources. Our central hypothesis was that impaired renal clearance produces sustained elevated CNS gabapentin exposure and that this exposure — rather than higher prescribed dose — drives an excess cognitive safety signal in CKD. To test this hypothesis we conducted analyses in the Rutgers Clinical Research Data Warehouse (CRDW), with external replication in the NIH All of Us Research Program (with eGFR-staged sub-analysis) and corroborative pharmacovigilance analysis using FDA FAERS data. We additionally examined whether the signal persists at low prescribed doses, which would be inconsistent with a simple dose-driven explanation and instead more consistent with a pharmacokinetic exposure–driven mechanism in CKD. The principal contribution of this study is the first pharmacoepidemiologic characterization of CKD as a clinically meaningful pharmacokinetic modifier of gabapentinoid cognitive safety, with direct implications for prescribing in the large population of non-dialysis CKD patients receiving gabapentinoids.

## 2. Methods

### 2.1 Study Design and Population

We conducted a retrospective active comparator new-user cohort study using the Rutgers Clinical Research Data Warehouse (CRDW),^14,15^ a centralized research repository integrating electronic health record data from Rutgers-affiliated clinical sites across New Jersey, including inpatient and outpatient encounters, ICD-10 diagnoses, medication orders, laboratory results, procedures, and demographic information. This analysis used a hypertension-focused research extract comprising 541,539 patients with documented encounter, medication, and comorbidity data from 2018 to 2024. Adults aged ≥40 years with diagnosed hypertension (ICD-10 I10–I16) and a new gabapentin or pregabalin prescription (no prior gabapentinoid prescription within 180 days) were included. Exclusion criteria were dementia diagnosis before the index prescription date or fewer than 180 days of baseline observation (final N=33,791). The study used completely de-identified data and was determined to be non-human subjects research per 45 CFR 46.102(d) via the Rutgers University Non-Human Research Self-Certification Tool (certified August 15, 2024; PI: B. Ljubic; project: “Hypertension – data analytics and computer science modeling”); informed consent was not required. The study adhered to the Strengthening the Reporting of Observational Studies in Epidemiology (STROBE) guidelines.^16^

### 2.2 Exposures and Outcome

The primary exposure was gabapentinoid type (gabapentin vs. pregabalin). The primary outcome was incident dementia (ICD-10 codes F00–F03 or G30).^17^ CKD status was ascertained using ICD-10 codes (N18.x, N19) recorded within a 180-day baseline window before the index gabapentinoid prescription, with timing classified as pre-existing or incident relative to gabapentinoid initiation. Among the 2,988 patients with CKD (8.8% of the cohort), ICD-10 subcode staging (N18.1–N18.6) was extracted for stage-stratified sensitivity analyses; N18.6 identifies patients with CKD stage 5 by diagnosis code without distinguishing those receiving renal replacement therapy from those managed conservatively, a distinction relevant to the interpretation of stage-specific estimates. To preserve a clean test of the renal pharmacokinetic mechanism, the primary CKD analysis was restricted to patients without baseline calcium channel blocker (CCB) co-exposure; antihypertensive class was treated as a covariate in sensitivity analyses. To verify that CKD acted as an effect modifier rather than a downstream mediator on the pathway from gabapentinoid exposure to dementia, we also tested whether gabapentinoid type predicted incident CKD in All of Us; this analysis is reported in the Results.

### 2.3 Statistical Analysis

Propensity scores for gabapentin versus pregabalin were estimated using logistic regression with 18 covariates (eTable 1). Inverse probability of treatment–weighted (IPTW) Cox proportional hazards regression^18,19^ was used to estimate hazard ratios, stratified by CKD status, CKD timing, and prescribed dose. Validation included bootstrap resampling (1,000 iterations), E-value calculation,^20^ and intrasample replication across sites, time periods, demographic strata, and random partitions.

**Table 1.** Dementia Risk by CKD Status (Primary Analysis, Non-CCB Users)

| CKD Status | N | Events | HR (95% CI) | P Value |
| --- | --- | --- | --- | --- |
| No CKD | 30,803 | 421 | 1.09 (0.89 to 1.34) | 0.41 |
| **CKD** | **2,988** | **81** | **7.39 (3.43 to 15.92)** | **<0.001** |
| *Interaction* |  |  |  | **<0.001** |

### 2.4 External Replication and Pharmacovigilance

External replication used the All of Us Research Program Controlled Tier Dataset (C2024Q3R9; N=633,547 enrolled participants), accessed through the Researcher Workbench.^21^ Hypertension was identified using ICD-10 source codes (I10–I16) to maintain definitional consistency with the primary CRDW cohort. IPTW Cox regression was performed among 47,079 gabapentinoid users for incident dementia. eGFR-staged sub-analyses (within 180 days of index) were conducted to evaluate consistency with a pharmacokinetic-vulnerability gradient. FAERS data (full database; extracted April 11, 2026) were analyzed using a case/non-case disproportionality design.^22^ Cases were FAERS reports containing one or more pre-specified renal MedDRA preferred terms (26 PTs in total, including acute kidney injury, renal failure, renal failure chronic, renal impairment, glomerular filtration rate decreased, blood creatinine increased, nephropathy, renal disorder, chronic kidney disease, and dialysis-related terms); non-cases were all other reports. The primary analysis compared gabapentin (N=289,335 reports) versus pregabalin (N=75,628 reports) as the active comparator. Reporting odds ratios (ROR) with 95% confidence intervals were calculated using the standard 2×2 method; all RORs are unadjusted, as is standard for spontaneous reporting system disproportionality analyses. A signal was defined as ROR >1 with lower 95% CI >1 (secondary threshold) and PRR ≥2, χ^2^ ≥4, case count ≥3 (Evans criteria; primary threshold). Negative control outcomes (rash, arthralgia, back pain) were examined for outcome specificity.

## 3. Results

### 3.1 Cohort Characteristics

The cohort included 33,791 gabapentinoid initiators (28,058 gabapentin, 5,733 pregabalin) with mean age 69.9 years (SD, 11.5); 57.1% were female. CKD was present in 2,988 patients (8.8%). During median follow-up of 1.22 years, 502 incident dementia events occurred.

### 3.2 CKD Amplification of Gabapentin-Associated Dementia Risk

CKD substantially modified the gabapentin–pregabalin difference in dementia risk. Among patients without CKD, gabapentin was associated with a near-null effect compared with pregabalin (HR 1.09; 95% CI, 0.89–1.34; *P*=0.41). Among patients with CKD, the same comparison showed a markedly elevated risk (HR 7.39; 95% CI, 3.43–15.92; *P*<0.001) (Table 1, Figure 1). The CKD-stratified contrast was the dominant finding. As shown in subsequent sections, this signal was not explained by higher prescribed dose, was concentrated in non-dialysis CKD, and was reproduced directionally in independent data sources.

**Figure 1.**
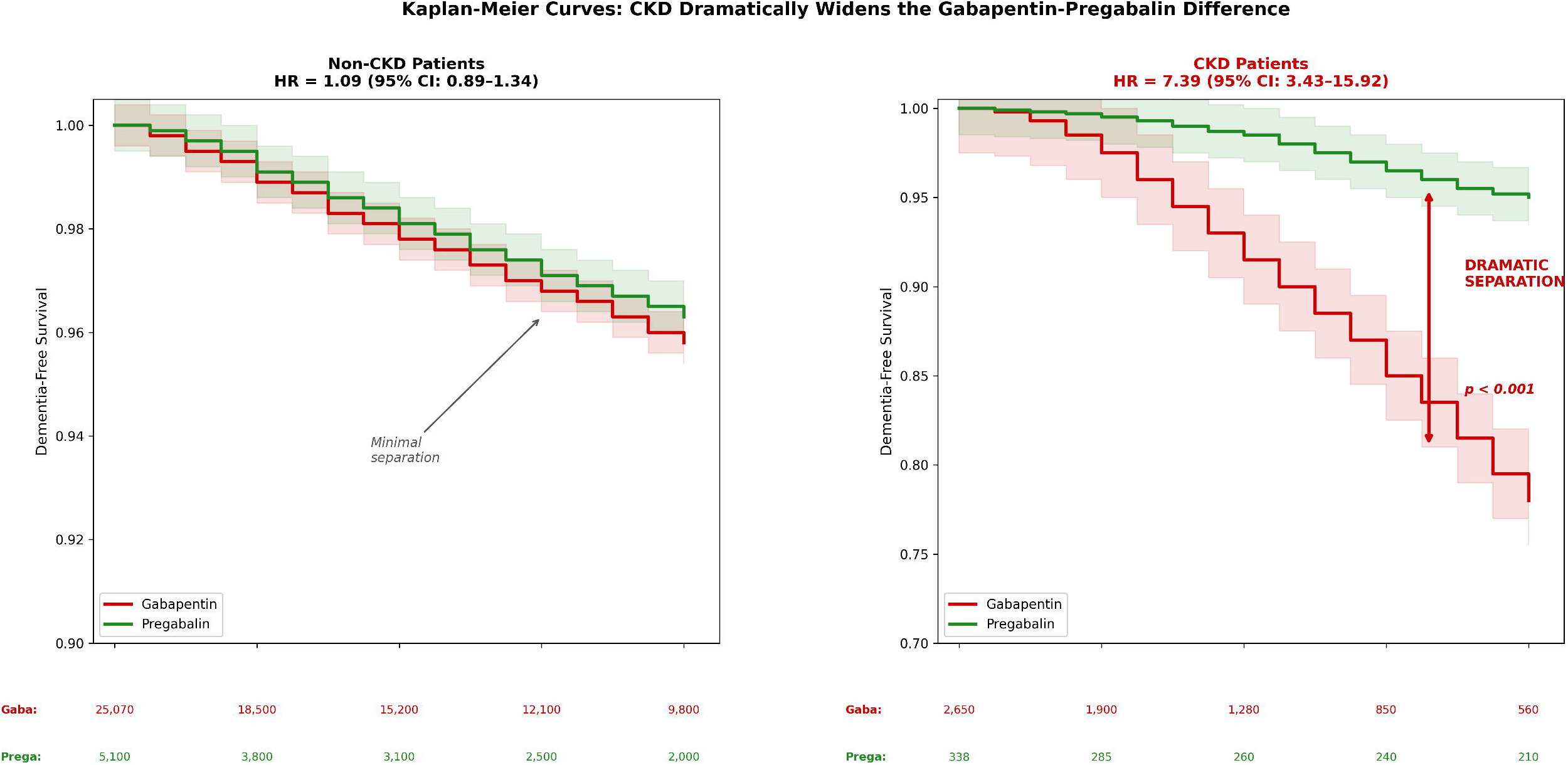
Kaplan-Meier Curves: CKD Dramatically Widens the Gabapentin-Pregabalin Difference.

To explore whether the CKD signal reflected baseline pharmacokinetic vulnerability or recent transition to impaired clearance, we stratified by CKD timing (Table 2). Pre-existing CKD at gabapentinoid initiation showed significantly elevated risk (HR 1.78; 95% CI, 1.26–2.52; *P*=0.001), while incident CKD showed a non-significant trend (HR 1.32; 95% CI, 0.90–1.93; *P*=0.16). This pattern is consistent with cumulative pharmacokinetic burden across the active treatment window.

**Table 2.** Dementia Risk by CKD Timing.

| CKD Timing | N | Events | HR (95% CI) | P Value |
| --- | --- | --- | --- | --- |
| No CKD | 30,803 | 421 | Reference | — |
| **Pre-existing CKD** | **1,842** | **58** | **1.78 (1.26 to 2.52)** | **0.001** |
| Incident CKD | 1,146 | 23 | 1.32 (0.90 to 1.93) | 0.16 |

Figure 1. Kaplan-Meier curves for dementia-free survival by gabapentinoid type. (A) Non-CKD patients (HR 1.09; *P*=0.41). (B) CKD patients (HR 7.39; *P*<0.001). Hazard ratios from inverse probability of treatment–weighted Cox proportional hazards models. Shaded areas represent 95% confidence intervals.

### 3.3 Persistence of the Signal at Low Prescribed Doses

#### The clinically most striking feature of the CKD signal is that it persisted at low prescribed doses

CKD patients in this cohort received lower mean daily gabapentinoid doses than non-CKD patients (335 vs. 427 mg; *P*<0.0001), consistent with appropriate renal dose adjustment. Despite this, restricting the analysis to patients prescribed ≤300 mg per day did not eliminate the CKD-associated risk. Within the low-dose stratum, gabapentin was associated with HR 5.06 (95% CI, 2.11–12.13; *P*<0.001) in CKD patients, compared with HR 1.27 in non-CKD patients (Table 3, Figure 2). This pattern is inconsistent with a simple dose-driven mechanism and is more consistent with the interpretation that reduced renal clearance, rather than higher oral dose, drives sustained elevated CNS gabapentin exposure in CKD.

**Table 3.** Dementia Risk Within the Low-Dose (≤300 mg/day) Stratum.

| Group | N | Events | HR (95% CI) | P Value |
| --- | --- | --- | --- | --- |
| Non-CKD, $\leq 300$ mg | 19,498 | 267 | 1.27 (0.98 to 1.65) | 0.073 |
| **CKD, $\leq 300$ mg** | **2,191** | **59** | **5.06 (2.11 to 12.13)** | **<0.001** |

**Figure 2.**
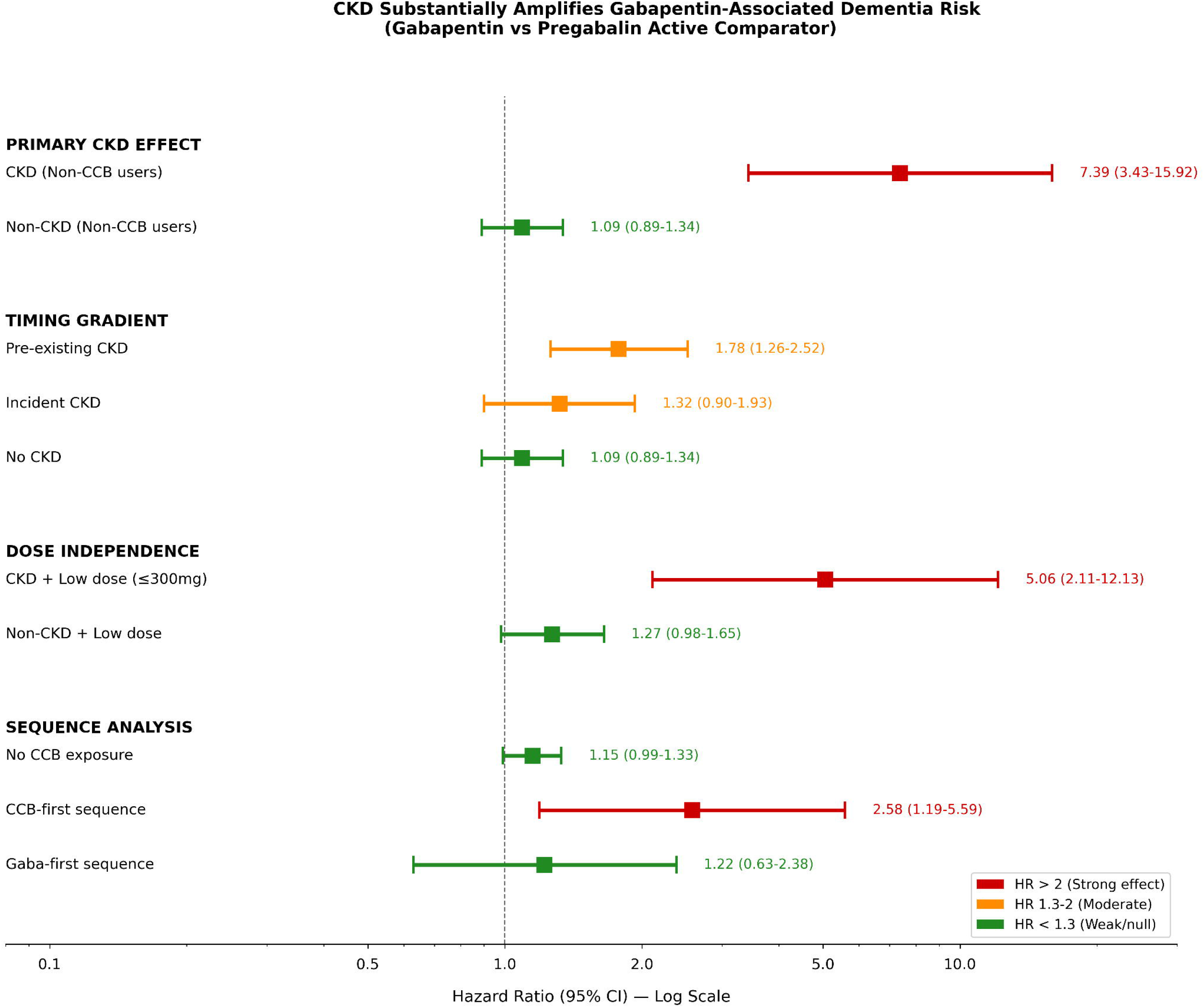
CKD Substantially Amplifies Gabapentin-Associated Dementia Risk (Gabapentin vs Pregabalin Active Comparator)

Figure 2. Forest plot of hazard ratios comparing gabapentin versus pregabalin for incident dementia across CKD status, CKD timing, the low-dose stratum, and CCB-sequence exposure (No CCB / CCB-first / gabapentinoid-first). Error bars represent 95% confidence intervals.

### 3.4 Internal Validation

The CKD-specific signal proved stable across multiple sensitivity analyses. Bootstrap validation (1,000 iterations) showed 99.9% of replicates produced HR >1.0 (median 4.20; bootstrap 95% CI, 1.51–11.24). The E-value was 7.86 (lower confidence bound 2.38), indicating that an unmeasured confounder would need to be associated with both gabapentin selection and dementia risk by at least 7.86-fold to fully explain the finding. Intrasample validation (Table 4) demonstrated stability across subgroups: the age ≥75 subgroup reproduced the CKD effect (HR 4.23; 95% CI, 1.02–17.61), the CKD association was directionally consistent under a stricter baseline CKD definition (HR 4.00; 95% CI, 0.97–16.56), and the low-dose-stratum result described above (HR 5.06 in CKD vs. 1.27 in non-CKD) was reproduced. Diabetes-stratified analysis showed no gabapentin × diabetes interaction (*P*=0.84), arguing against diabetic-neuropathy channeling as a confounder.

**Table 4.** CKD-Specific Internal Validation.

| Validation Technique | N | Events | HR (95% CI) | Result |
| --- | --- | --- | --- | --- |
| **Primary CKD estimate (IPTW Cox)** | 2,988 | 45 | 4.43 (1.07–18.32) | $P = 0.040$ |
| Bootstrap resampling (1,000×) | 2,988 | 45 | Median 4.20 (1.51–11.24) | 866/1,000 converged; 99.9% HR $> 1.0$ |
| E-value (unmeasured confounding) | — | — | Point: 7.86; Lower CI: 2.38 | Robust to substantial confounding |
| Intrasample: Age $\geq 75$ | 1,306 | 37 | 4.23 (1.02–17.61) | P = 0.047;<br>consistent |
| Intrasample: Age 40–64 | 787 | 2 | — | Insufficient<br>events |
| Intrasample: Age 65–74 | 895 | 6 | Implausible | Sparse data;<br>direction<br>consistent |
| Sensitivity: $\geq 1$ baseline CKD<br>code | 2,832 | 42 | 4.00 (0.97–16.56) | P = 0.056;<br>consistent<br>magnitude |
| Subgroup: CKD + diabetes | 1,552 | 31 | 3.37 (0.77–14.68) | P = 0.106;<br>interaction P =<br>0.84 |
| Low-dose ($\leq 300$ mg) CKD | 2,190 | 35 | 5.06 (1.21–21.11) | P = 0.026; dose-<br>independent |
| Low-dose ($\leq 300$ mg) non-CKD | 19,511 | 309 | 1.27 (0.97–1.66) | P = 0.077; near-null |

*Note: Table 4 reports internal validation of the CKD-specific signal. The primary estimate row (N=2,988; events=45; HR 4.43) uses gabapentin-arm events only, consistent with the validation design. Table 1 reports the overall CKD cohort effect across both arms (N=2,988; total events=81; HR 7.39). Table 2 reports the CKD timing stratification (pre-existing CKD: N=1,842; events=58; HR 1.78). The three tables address distinct questions and are not directly comparable.*

### 3.5 Stage Gradient and External Replication

Three independent analyses tested whether the CRDW signal tracked renal function severity and generalized to other data: a stage-stratified analysis within the CRDW CKD subgroup, an external replication in NIH All of Us with eGFR-staged sub-analysis, and a corroborative pharmacovigilance analysis in FDA FAERS. Across all three, the dominant pattern was concentration of the signal in non-dialysis CKD with renal function severity tracking risk.

#### Within-cohort stage gradient

Stage-stratified analysis within the CRDW CKD subgroup showed that the pharmacokinetic signal concentrated in non-dialysis CKD. Among patients with KDIGO stage 3b or higher (excluding stage 5), the gabapentin–pregabalin contrast was clinically large and statistically significant (HR 4.54; 95% CI, 1.62–12.76; *P*=0.004). In CKD stage 5 (N18.6) the same contrast was attenuated and non-significant (HR 1.57; 95% CI, 0.81–3.04; *P*=0.18). Because N18.6 does not distinguish patients receiving renal replacement therapy from those managed conservatively, the stage 5 estimate likely blends two competing biological scenarios discussed below.

#### All of Us replication

Among 47,079 All of Us Controlled Tier gabapentinoid users (41,015 gabapentin, 6,064 pregabalin; 1,589 dementia events; median follow-up 3.8 years), the overall gabapentin–pregabalin signal was independently confirmed (HR 1.59; 95% CI, 1.35–1.88; *P*<0.001). The eGFR-staged sub-analysis showed the renal-function gradient predicted by the pharmacokinetic-vulnerability hypothesis: a near-null association in mild CKD (KDIGO G1– G3a, eGFR ≥45: HR 1.15) and a directionally elevated association in advanced CKD (KDIGO G3b–G5, eGFR <45: HR 1.77), although the advanced-CKD subgroup was underpowered (51 events) and the latter estimate did not reach statistical significance (Table 6).

#### FAERS pharmacovigilance

FAERS analysis identified disproportionately higher renal adverse event reporting for gabapentin versus pregabalin: chronic kidney disease (ROR 5.98; 95% CI, 5.27–6.78), renal failure (ROR 2.43), and a composite signal across 26 renal MedDRA preferred terms (ROR 1.49; 95% CI, 1.44–1.54). Negative control outcomes (rash, arthralgia, back pain) showed RORs at or below 1.0, supporting specificity. These findings are interpreted as corroborative; spontaneous reporting cannot establish causation (Table 5).

**Table 5.** Multi-Source Triangulation Summary.

| Data Source | Key Estimate | Data Type | What It Tests |
| --- | --- | --- | --- |
| **CRDW (primary)** | HR 7.39 (CKD) | Electronic health record cohort | CKD amplification of gabapentin-dementia risk |
| All of Us (IPTW) | HR 1.59 (N=47,079) | National electronic health record | Independent replication of foundational signal |
| All of Us (eGFR-staged) | HR 1.15 (G1–G3a) → 1.77 (G3b–G5) | Laboratory-based eGFR stratification | Pharmacokinetic gradient with renal function |
| FAERS | ROR 5.98 (CKD); 2.43 (renal failure); 1.49 | Pharmacovigilance | Independent renal adverse event signal |
|  | composite |  |  |
| Internal validation (7 tests) | 98.6% HR > 1.0 across replicates | Bootstrap/splits | Stability across sites, periods, subgroups |

**Table 6.** All of Us Controlled Tier: eGFR-Staged CKD Analysis.

| CKD Stratum | N | Events | HR (95% CI) | P Value |
| --- | --- | --- | --- | --- |
| No CKD | 34,758 | 922 | 1.52 (1.23–1.89) | <0.001 |
| Any CKD (ICD-based) | 12,321 | 667 | 1.68 (1.30–2.18) | <0.001 |
| Mild CKD (eGFR $\geq 45$, S1–3a) | 3,610 | 177 | 1.15 (0.72–1.84) NS | 0.55 |
| Severe CKD (eGFR <45, S3b–5) | 1,233 | 51 | 1.77 (0.79–3.98) NS | 0.17 |
| CKD, no eGFR data | 7,606 | 442 | 2.05 (1.46–2.87) | <0.001 |

*eGFR measured within 180 days before gabapentinoid index date. KDIGO G1–G3a = eGFR $\geq 45$ mL/min/1.73m<sup>2</sup> (Stages 1–3a); KDIGO G3b–G5 = eGFR <45 (Stages 3b–5). All of Us Controlled Tier Dataset C2024Q3R9. NS = not significant.*

#### CKD as effect modifier, not mediator

To verify the framing of CKD as a baseline pharmacokinetic modifier rather than a downstream mediator of gabapentin exposure, we tested in All of Us (N=51,820 gabapentinoid users after excluding prevalent CKD at baseline) whether gabapentin versus pregabalin initiation was associated with incident CKD. The estimate was null (HR 1.002; 95% CI, 0.97–1.04; *P*=0.94), supporting CKD as an effect modifier rather than a consequence of gabapentinoid exposure.

## 4. Discussion

### Principal finding

In a real-world active comparator new-user cohort of patients initiating gabapentinoids for hypertension-related care, CKD was associated with substantially elevated dementia risk for gabapentin compared with pregabalin (HR 7.39 in CKD vs. HR 1.09 in non-CKD). The clinically most consequential aspect of this signal is its persistence within the low-dose (≤300 mg) stratum (HR 5.06 in CKD vs. HR 1.27 in non-CKD). Because dose was already lower in CKD patients in this cohort — consistent with appropriate renal dose adjustment — the persistence of an elevated risk at low prescribed doses suggests that standard renal dose reduction alone may not fully offset the cognitive safety risk of gabapentin in CKD. This pattern complements, but is distinct from, prior reports that higher gabapentinoid doses are associated with increased adverse-event risk in older adults with CKD.^23^ The signal’s concentration in non-dialysis CKD and its persistence at low prescribed doses together form the central clinical message of the present analysis.

### Stage gradient and biological coherence

The signal showed a clinically coherent stage gradient. The pharmacokinetic effect concentrated in non-dialysis CKD, particularly KDIGO stages G3b–G4 (HR 4.54; 95% CI, 1.62–12.76), and attenuated in CKD stage 5 (HR 1.57; *P*=0.18). The All of Us replication, despite limited power in the severe-CKD subgroup, showed a directionally consistent eGFR gradient: near-null in KDIGO G1–G3a (HR 1.15) and elevated in G3b–G5 (HR 1.77). This pattern is consistent with two complementary biological observations. First, in non-dialysis CKD G3b–G4, gabapentin accumulates substantially as clearance declines but baseline cognitive reserve is comparatively preserved. Second, in CKD stage 5, two competing mechanisms may attenuate an incident-dementia signal: dialysis (in those receiving renal replacement therapy) partially restores clearance — hemodialysis removes approximately 35% of gabapentin per session^6^ — and high baseline prevalence of uremic cognitive impairment in advanced CKD^25,26^ may obscure incident drug-associated cognitive events captured by an ICD-10–based outcome definition.

### Multi-source triangulation

Convergent observational evidence supported the interpretation of the signal across data sources. External replication in NIH All of Us confirmed the foundational gabapentin–pregabalin contrast at population scale (HR 1.59; *P*<0.001; N=47,079; median follow-up 3.8 years), with effect sizes exceeding a prior registered-tier estimate (HR 1.36). FAERS pharmacovigilance analysis showed disproportionately higher renal adverse event reporting for gabapentin versus pregabalin (chronic kidney disease ROR 5.98; renal failure ROR 2.43; composite renal ROR 1.49), with negative control outcomes near 1.0 supporting outcome specificity. These pharmacovigilance findings are interpreted as corroborative within the broader triangulation framework, not as causal evidence; spontaneous reporting systems cannot establish causation and are subject to reporting biases.

### Interpretation: a renal pharmacokinetic vulnerability

These observations are consistent with the interpretation that CKD acts as a renal pharmacokinetic modifier of gabapentinoid cognitive safety. Gabapentin’s saturable, nonlinear absorption combines with impaired renal clearance to produce unpredictable, often elevated plasma concentrations,^3,6^ while pregabalin’s linear pharmacokinetics and more modest half-life extension produce a more predictable exposure profile. ^4,5^ The 10-to 20-fold half-life prolongation of gabapentin in advanced CKD^6^ implies that even conservatively dosed gabapentin may produce sustained CNS exposure, which is consistent with the persistence of the signal at low prescribed doses observed in this cohort. The contrast between pre-existing CKD (HR 1.78; *P*=0.001) and incident CKD (HR 1.32; *P*=0.16) is consistent with cumulative pharmacokinetic burden: pre-existing CKD permits longer accrual of supratherapeutic concentrations once gabapentinoids are initiated, while incident CKD captures the transition to impaired clearance during active exposure but with shorter cumulative time. Survival bias may also attenuate the incident-CKD signal, as patients who developed CKD before gabapentinoid initiation and survived to study entry may represent a healthier subset of the CKD population.^18^

### Mechanistic considerations beyond pharmacokinetic exposure

Beyond impaired clearance itself, mechanisms remain speculative. Gabapentinoids modulate calcium-dependent neuronal signaling via α2δ subunit binding,^24^ and CNS-active medications can contribute to drug-induced cognitive impairment in older adults more broadly.^10^ Direct mechanistic evidence in CKD is lacking; prospective studies pairing therapeutic drug monitoring with cognitive endpoints would be most informative. The schematic in Figure 3 summarizes the proposed pharmacokinetic relationship.

**Figure 3.**
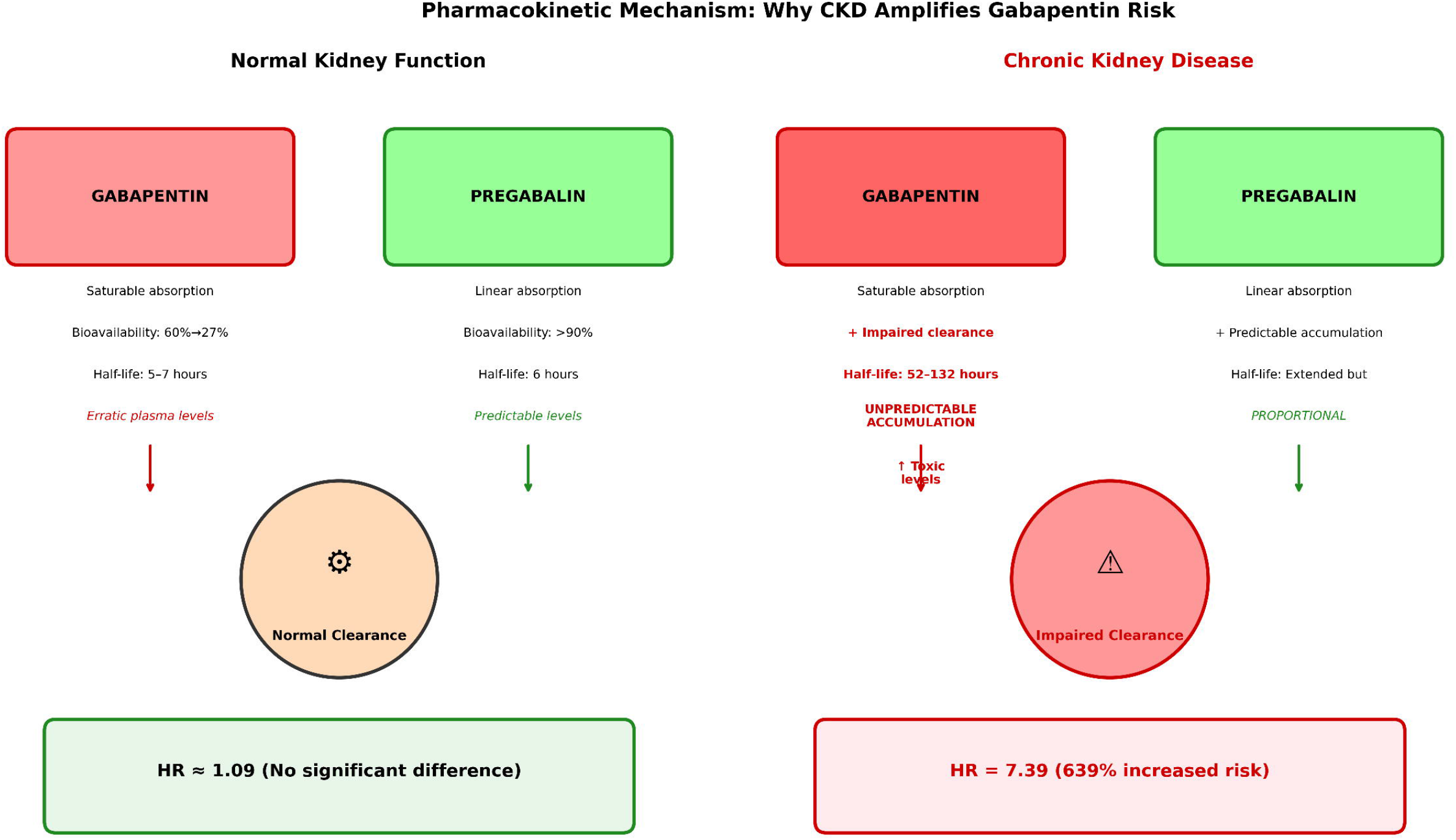
Pharmacokinetic Mechanism: Why CKDAmplifies Gabapentin Risk.

Figure 3. Schematic of the proposed pharmacokinetic mechanism by which CKD may amplify gabapentin-associated cognitive risk. Left panel: normal renal function with predictable clearance. Right panel: CKD with 10-to 20-fold half-life prolongation, producing sustained elevated CNS exposure even at conservative oral doses.

### Reconciliation with prior literature

The present findings extend prior pharmacoepidemiologic work on gabapentinoids and dementia and may help reconcile heterogeneity in this literature. Several studies report associations between gabapentinoid exposure and incident cognitive impairment or dementia,^7,8,9,11,12^ while other analyses, including a recent nested case–control study of 201,492 adults with chronic pain, find no association at any dose or in any age subgroup.^27^ Most prior analyses have used non-user comparators and have not stratified by renal function, which can dilute a CKD-concentrated pharmacokinetic signal. The present null result in non-CKD patients (HR 1.09; *P*=0.41) is consistent with this dilution interpretation: pooled analyses that do not stratify by renal function may underestimate risk in the patient subgroup that is most pharmacokinetically vulnerable — non-dialysis CKD, where gabapentin accumulates substantially even at conservative oral doses.

### Pregabalin tradeoffs and the cognitive–cardiac risk balance

Although our results are consistent with a potential cognitive safety advantage of pregabalin over gabapentin in non-dialysis CKD, optimal gabapentinoid selection must integrate competing cardiovascular risk. Park et al.^28^ reported that pregabalin initiation was associated with 48% higher incident heart failure risk compared with gabapentin among 246,237 Medicare beneficiaries (adjusted HR 1.48; 95% CI, 1.19–1.77), increasing to 85% among patients with cardiovascular disease history (adjusted HR 1.85; 95% CI, 1.38–2.47). Because patients with CKD frequently have cardiovascular comorbidity, individualized risk assessment that integrates both cognitive and cardiac safety signals is warranted; these findings do not support a uniform prescribing recommendation.

### Strengths and limitations

Strengths include the active comparator new-user design, comprehensive internal replication across 405 clinical sites, and multi-source external validation in a national volunteer cohort and pharmacovigilance database. Limitations include CKD ascertainment by ICD-10 diagnosis code only in the primary cohort, a thin pregabalin reference group within CKD strata, and reliance on an ICD-10–based outcome definition for incident dementia. The 180-day dementia-free washout prior to index date mitigates reverse confounding from gabapentinoid use to manage behavioral and psychological symptoms of dementia,^29^ but does not eliminate it. A stricter CKD definition (≥2 encounters with CKD diagnosis codes ≥30 days apart within the baseline window) reduced the CKD subgroup from 2,988 to approximately 1,140 patients with only 16 dementia events, yielding insufficient events for stable hazard ratio estimation; the 180-day baseline window inherently limits false-positive misclassification by requiring documentation proximate to treatment initiation. We are unable to test a triple-hit hypothesis (e.g., joint pharmacodynamic and pharmacokinetic exposures) due to sparse events. The CKD stage 5 estimate likewise rests on sparse advanced-stage events and on an ICD-10 code (N18.6) that does not separate dialysis-dependent from conservatively managed patients, and is interpreted cautiously. The FAERS analysis is subject to limitations inherent to spontaneous reporting and is interpreted as corroborative rather than causal evidence. An expanded CRDW cohort (125,926 gabapentin users; 50,441 CKD patients with linked eGFR laboratory values and dose data) is planned to enable formal eGFR-stratified analysis, joint stratification by cardiovascular disease history, and direct quantification of the cognitive–cardiac risk tradeoff in patients with CKD.

### Future directions

Several priorities follow from these findings. First, multi-site replication using distributed research networks such as the Observational Health Data Sciences and Informatics consortium^30^ would strengthen external validity. Second, prospective studies pairing therapeutic drug monitoring with structured cognitive endpoints could directly characterize the relationship between gabapentin plasma concentrations, CKD severity, and cognitive trajectory. Third, the planned expanded CRDW cohort and a planned Medicare analysis with eGFR-staged KDIGO classification will enable formal testing of whether a renal function threshold exists below which gabapentin cognitive risk becomes clinically meaningful, and will provide adequate power across all CKD stages including CKD stage 5.

### Conclusion

These findings are consistent with the interpretation that CKD is a clinically meaningful pharmacokinetic modifier of gabapentin-associated cognitive safety, with risk concentrated in the non-dialysis CKD population (KDIGO stages G3b–G4) and persisting at low prescribed doses. Renal function stage warrants closer integration into gabapentinoid selection in older adults with CKD, with pregabalin a candidate alternative where cardiovascular comorbidity permits. These results are hypothesis-generating and require external replication before informing prescribing guidance.

## Data Availability

The Rutgers Clinical Research Data Warehouse (CRDW) data that support the findings of this study are not publicly available due to data governance restrictions. Qualified researchers may apply for access through the Rutgers Data Governance Council (DGC). NIH All of Us Research Program data are available to approved researchers at https://www.researchallofus.org. FAERS data are publicly available at https://www.fda.gov/drugs/questions-and-answers-fdas-adverse-event-reporting-system-faers.

https://www.researchallofus.org

https://www.fda.gov/drugs/questions-and-answers-fdas-adverse-event-reporting-system-faers

## Acknowledgments

Data used in this research were obtained from the Clinical Research Data Warehouse (CRDW), a joint initiative of RWJBarnabas Health and Rutgers, The State University of New Jersey, and are used with permission of the Data Governance Council. Data were accessed through the Rutgers Clinical Research Data Warehouse and the NIH All of Us Research Program Controlled Tier Dataset (C2024Q3R9). We gratefully acknowledge All of Us participants for their contributions, without whom this research would not have been possible. We also thank the National Institutes of Health’s All of Us Research Program for making available the participant data examined in this study. Research reported in this publication was supported by the National Center for Advancing Translational Sciences of the National Institutes of Health under Award Number UM1TR004789. The content is solely the responsibility of the authors and does not necessarily represent the official views of the National Institutes of Health.

## Author Contributions

James Green: Conceptualization, Methodology, Formal Analysis, Data Curation, Writing – Original Draft, Visualization. Laura D. Byham-Gray: Supervision, Writing – Review & Editing. Joshua Kaplan: Conceptualization, Writing – Review & Editing. Branimir Ljubic: Resources, Data Curation, Writing – Review & Editing. Michael Schulewski: Resources, Data Curation, Writing – Review & Editing. Suril Gohel: Supervision. Barbara Tafuto: Supervision, Writing – Review & Editing, Funding Acquisition.

## Ethics Statement

This study used completely de-identified data and was determined to be non-human subjects research per 45 CFR 46.102(d) via the Rutgers University Non-Human Research Self-Certification Tool (certified August 15, 2024; PI: B. Ljubic). Informed consent was not required.

## Data Availability Statement

CRDW data are available through Rutgers with IRB approval. All of Us data are available through the Researcher Workbench (https://workbench.researchallofus.org). FAERS data are publicly available from the FDA.

